# Atrial Fibrillation Polygenic Risk Score (AF-PRS) Predicts Non-Ischemic Cardiomyopathy: A Single-Center Retrospective Cohort Study of 16,801 Individuals

**DOI:** 10.64898/2026.03.20.26348938

**Authors:** Mahmoud Alsaiqali, Blerina Asllanaj, Victor Florea, Logan Johnke, Whitnee Otto, Max Weaver, Shubhi Bartaraia, Shaopeng Gu, Jerome I. Rotter, Xiuqing Guo, Jingyi Tan, Henry J. Lin, Colette Free, Holly Takkinen, Eric Larson, Catherine Hajek, Adam Stys, Natalia Baran, Tomasz Stys

**Affiliations:** Sanford Cardiovascular Institute, University of South Dakota Sanford School of Medicine, Sioux Falls, SD; Sanford Health, Sioux Falls, SD; The Institute for Translational Genomics and Population Sciences, Department of Pediatrics, The Lundquist Institute for Biomedical Innovation at Harbor-UCLA Medical Center, Torrance, CA, USA; Helix LLC; Department of Internal Medicine, University of South Dakota Sanford School of Medicine, Sioux Falls, SD; Department of Hematology and Hematological Central laboratory, Inselspital Bern, Bern University Hospital, Bern, Switzerland

## Abstract

**Background:** Non-ischemic cardiomyopathy (NICM) represents a major cause of heart failure with limited tools for early risk stratification. Atrial fibrillation (AF) is a well-established contributor to cardiomyopathy but is often clinically silent in its early stages. The atrial fibrillation polygenic risk score (AF-PRS) reflects genetic susceptibility to AF and may identify individuals at risk for AF-related cardiomyopathy. We hypothesized that higher AF-PRS is associated with greater risk of NICM.

**Methods:** This was a retrospective cohort study of 16,801 individuals of European ancestry from the Sanford Biobank and Imagenetics program with genetic sequencing and longitudinal electronic health record data. AF-PRS was calculated using 315 genome-wide significant single-nucleotide polymorphisms with standard quality control. NICM was defined by International Classification of Diseases, 10th Revision, Clinical Modification codes, excluding ischemic etiologies. Cox regression models evaluated the association between AF-PRS and incident NICM, adjusting for age, sex, smoking status, body mass index (BMI), hypertension, and diabetes. AF-PRS was analyzed both as a quasi-continuous variable (15% quartile increments) and dichotomized at the 85th percentile. Sensitivity analyses assessed associations with all-cause cardiomyopathy and ischemic cardiomyopathy. Survival analysis was used to model time-to-event outcomes.

**Results:** Among all participants, 418 (2.5%) had NICM. 99% were Caucasian. NICM cases were older and more often male (both p<0.001) than those without a diagnosis. After multivariable adjustment for sex, smoking status, BMI, and hypertension, a linear AF-PRS (15% increments) was specifically predictive of increased hazard risk of NICM (HR = 1.09 [1.03, 1.15], p < 0.001).

**Conclusion:** These findings complement recent evidence of bidirectional genetic relationships between cardiomyopathy and AF, supporting comprehensive genetic risk assessment in cardiovascular disease prevention. Clinical implementation requires validation in diverse populations and prospective evaluation. Future research should investigate the mechanistic pathways linking AF-associated genetic variants to cardiomyopathy development and evaluate whether AF-PRS-guided screening improves clinical outcomes.

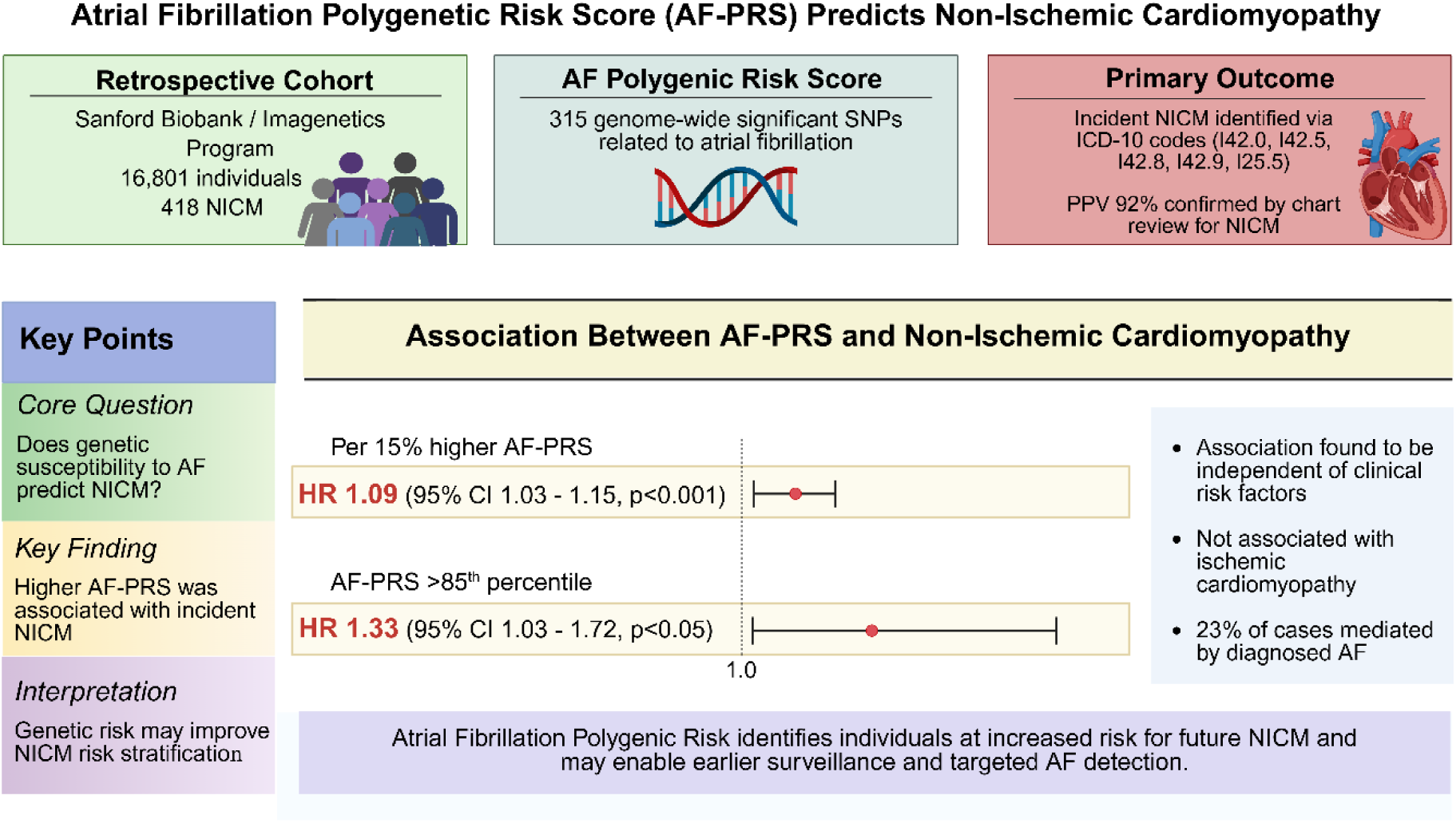

**Visual abstract: Summary of study design, exposure, outcome, key findings, and clinical implications.:** This graphical abstract provides a comprehensive overview of the study investigating the association between atrial fibrillation polygenic risk score (AF-PRS) and non-ischemic cardiomyopathy (NICM). Abbreviations: AF, atrial fibrillation; AF-PRS, atrial fibrillation polygenic risk score; CI, confidence interval; HR, hazard ratio; ICD-10, International Classification of Diseases, 10th Revision; ICM, ischemic cardiomyopathy; NICM, non-ischemic cardiomyopathy; PPV, positive predictive value; PRS, polygenic risk score; SNP, single nucleotide polymorphism. Created in BioRender. https://BioRender.com/ k3t1w68

## Introduction

Cardiomyopathy and heart failure (HF) represent major public health challenges with substantial morbidity, mortality, and healthcare costs. According to the most recent epidemiological data, approximately 6.7 million Americans over 20 years of age have heart failure, with projections indicating the number will increase to 8.7 million by 2030.^1^ In 2021, there were 1.2 million primary HF hospitalizations in the United States, and HF was a contributing cause in approximately 425,147 deaths, accounting for 45 % of cardiovascular deaths^1^. Updated 2024-2025 statistics indicate that age-adjusted prevalence of HF continues to rise, particularly among older adults and individuals with multiple comorbidities, with annual direct and indirect costs exceeding 50 billion dollars^1^. The five-year mortality rate for HF remains approximately 50%, underscoring the critical need for improved prevention and early detection strategies.^2^

Non-ischemic cardiomyopathy (NICM) accounts for approximately 40-50% of all heart failure cases and encompasses diverse etiologies including dilated cardiomyopathy, hypertrophic cardiomyopathy, restrictive cardiomyopathy, and arrhythmia-induced cardiomyopathy.^2^ Unlike ischemic cardiomyopathy, which results from coronary artery disease and myocardial infarction, NICM arises from genetic, inflammatory, toxic, or idiopathic causes.^3^ Early identification of individuals at risk for NICM could enable targeted surveillance, lifestyle modifications, and preventive therapies to delay or prevent disease onset. Traditional cardiovascular risk prediction models rely primarily on clinical factors such as age, sex, blood pressure, cholesterol levels, smoking status, and diabetes.^1^ While these models have proven valuable, they explain only a portion of cardiovascular disease risk, and substantial inter-individual variation remains unexplained^4^. Genetic factors contribute significantly to cardiovascular disease susceptibility, with heritability estimates ranging from 30% to 60% for various cardiovascular phenotypes.^4^ Thus, besides traditional risk factors, genetics can be additive in risk prediction.^4^ A single gene mutation can explain a minority of cardiovascular diseases; however, most diseases result from polygenetic mutations or variations that interact with environmental factors.^5^

Genome-wide association studies (GWAS) have identified thousands of common single-nucleotide variants (SNVs) that are associated with various cardiovascular diseases and risk factors.^6–8^ Polygenic risk scores (PRS) aggregate the effects of multiple genetic variants into a single quantitative measure of genetic predisposition.^5^ Each variant contributes a small effect, but collectively, they can substantially influence disease risk. ^9^ Different SNVs are combined in one score called a polygenic risk score (PRS). PRS have been developed for numerous cardiovascular conditions, including coronary artery disease (CAD), atrial fibrillation (AF), hypertension, and hyperlipidemia (HLP).^9,10^ Recent evidence demonstrates that PRS can identify individuals at substantially elevated cardiovascular risk, sometimes comparable to or exceeding the risk conferred by monogenic mutations.^9^ For example, individuals in the top decile of coronary artery disease PRS have approximately 3-fold increased risk compared to those in the middle quintile.^5^ Importantly, PRS provide risk information independent of traditional clinical risk factors and can identify high-risk individuals at younger ages, potentially enabling earlier intervention.^10^

Atrial fibrillation (AF) is the most common sustained cardiac arrhythmia, affecting approximately 6 million Americans, with prevalence projected to exceed 12 million by 2030.^2^ AF is a well-established contributor to cardiomyopathy through multiple mechanisms, including tachycardia-induced myocardial dysfunction, loss of atrial contribution to ventricular filling, irregular ventricular response, and thromboembolism, and is associated with a high incidence and prevalence of HF.^2,11^ Tachycardia-induced cardiomyopathy, also termed “arrhythmia-induced cardiomyopathy,” can develop when sustained rapid ventricular rates lead to progressive left ventricular dysfunction.^12,13^ Importantly, this form of cardiomyopathy is potentially reversible with rate control or rhythm restoration.^12^

A critical challenge in understanding the AF-cardiomyopathy relationship is that AF is frequently asymptomatic, particularly in its early stages. Studies indicate that approximately 40% of patients with AF are asymptomatic, and many individuals have undiagnosed “subclinical” AF detected only through prolonged monitoring or incidentally during medical evaluations.^14^ This silent AF may contribute to cardiomyopathy development before clinical diagnosis, representing a missed opportunity for early intervention.

The atrial fibrillation polygenic risk score (AF-PRS) integrates genetic variants robustly associated with AF susceptibility identified through large-scale GWAS involving hundreds of thousands individuals^15^. These studies have identified over 300 independent genetic loci associated with AF, implicating genes involved in cardiac development, ion channel function, structural remodeling, and transcriptional regulation.^15^ AF-PRS has been validated as a predictor of incident AF in multiple independent cohorts, with individuals in the highest PRS decile having 3-4 fold increased AF risk compared to those in the lowest decile.^9^ Emerging evidence suggests that genetic variants associated with AF may also influence cardiac structure and function beyond their effects on arrhythmia susceptibility.^15^ Several AF-associated genes, including those encoding transcription factors (PITX2, TBX5), ion channels (SCN5A, KCNH2), and structural proteins, play roles in cardiac development and myocardial function.^15^ These shared genetic pathways raise the possibility that AF-PRS may predict cardiomyopathy risk through mechanisms beyond AF itself.

Despite extensive research on genetic risk factors for specific cardiomyopathy subtypes (e.g., hypertrophic cardiomyopathy, dilated cardiomyopathy), no previous studies have examined whether AF-PRS predicts NICM risk in the general population. Previous attempts to develop PRS for heart failure have been limited by heterogeneous phenotype definitions, modest effect sizes, and lack of specificity for cardiomyopathy.^16,17^ Studies focusing on specific cardiomyopathy types with more precise phenotyping have shown more promising results.^18,19^

We hypothesized that AF-PRS would be associated with increased NICM risk through two potential mechanisms: (1) undiagnosed or subclinical AF leading to tachycardia-induced cardiomyopathy, and (2) shared genetic pathways affecting both cardiac electrophysiology and myocardial structure/function. We further hypothesized that this association would be specific to NICM rather than ischemic cardiomyopathy (ICM), as AF-related mechanisms are more relevant to non-ischemic etiologies.

The primary objective of this study was to evaluate the association between AF-PRS and incident NICM in a large cohort of individuals with genetic data and longitudinal electronic health records. Secondary objectives included: (1) assessing whether AF-PRS predicts NICM independent of traditional cardiovascular risk factors, (2) evaluating the specificity of the association by examining relationships with all-cause cardiomyopathy and ICM, and (3) exploring potential clinical implications for risk stratification and prevention.

## Methods

### Study population

This retrospective cohort study utilized data from the Sanford Biobank and Imagenetics program, which includes individuals who voluntarily provided biological specimens and consented to research use of their genetic and clinical data. The Sanford Health system serves populations primarily in North Dakota, South Dakota, Minnesota, and Nebraska. Eligible participants were 18 years or older at enrollment, had available genetic data from genome-wide genotyping, and had linked longitudinal electronic health record (EHR) data. The study was conducted under protocols approved by the Sanford Health Institutional Review Board (IRB), which waived requirements for individual informed consent and HIPAA authorization due to the use of de-identified data and minimal risk to participants.

The study cohort was restricted to individuals of European ancestry to minimize population stratification bias in genetic analyses, as the AF-PRS was derived from predominantly European ancestry GWAS. Ancestry was determined using principal components analysis of genome-wide genotype data, with individuals clustering with European reference populations (1000 Genomes Project) included in the analysis.

### Genotyping, Quality Control and Genotype Imputation

Participants were genotyped using the Illumina Global Screening Array (GSA) version 1, which assays approximately 700,000 genetic variants across the genome. Standard quality control procedures were applied at both the sample and variant levels. Samples were excluded if they had: (1) genotyping call rate < 95%, (2) excess heterozygosity (> 3 standard deviations from the mean), (3) sex discordance between genetic and reported sex, or (4) cryptic relatedness (identity-by-descent > 0.185, indicating second-degree relatives or closer). Genetic variants were excluded if they had: (1) call rate < 95%, (2) minor allele frequency < 0.01, or (3) significant deviation from Hardy-Weinberg equilibrium (p < 1×10⁻⁶).

To expand coverage beyond directly genotyped variants, genotype imputation was performed using the Michigan Imputation Server with the Haplotype Reference Consortium (HRC) reference panel (version r1.1). Pre-imputation quality control included strand alignment and allele frequency checks. Post-imputation, variants were retained only if they had imputation quality score (r²) > 0.3, ensuring adequate imputation accuracy. This threshold balances the inclusion of additional variants while maintaining data quality.

### Atrial Fibrillation Polygenic Risk Score Calculation

AF-PRS was calculated using 315 genome-wide significant SNPs identified from three large-scale GWAS of atrial fibrillation.^20–22^ These studies collectively analyzed over 1 million individuals and identified robust genetic associations with AF.

These SNPs served as the foundation for our research. To ensure that our genetic variant selection included only independent SNPs, we performed linkage disequilibrium (LD) pruning. LD refers to the non-random association of alleles at different loci on the same chromosome. By pruning, we removed SNPs that were in high LD with each other (r2 < 0.2), eliminating redundancy in our dataset. This step ensured that our analysis focused on distinct and informative genetic variants. In some cases, specific SNPs exhibit palindromic behavior, meaning that their alleles have the same frequency. This symmetry can create challenges during subsequent analysis. To address this issue, we replaced palindromic SNPs with proxies that had allele frequencies ranging from 0.35 to 0.65. These proxies were selected based on a high level of LD (r2 > 0.8) and D’ (a measure of linkage disequilibrium). By using proxies, we maintained the genetic diversity needed for accurate analysis while overcoming the limitations posed by palindromic SNPs.

To ensure comprehensive genetic variant selection, we utilized imputed data. Imputation is a statistical technique that allows us to infer the genotypes of ungenotyped SNPs based on patterns observed in nearby genotyped SNPs. We only included variants with a high imputation quality score (r2 > 0.3) in our analysis. By doing so, we expanded the scope of our study, capturing a wider range of genetic variations.

We calculated the weighted continuous genetics risk scores using a formula that considered several key parameters. For each subject, the polygenic score (PRS) was computed, incorporating the posterior probability (dosage) of being a heterozygous or homozygous effect allele carrier after imputations for each variant. The number of independent variants (κ) in the polygenic score and the weight for each variant obtained from GWAS summary statistics were also considered. High AF-PRS was classified as >85th percentile, and low to intermediate AF-PRS was ≤ 85th percentile.

To ensure comparability across cohorts, we standardized the risk scores for each of the five cohorts (RS-I, RS-II, RS-III, MESA, and Sanford Health). The standardization involved transforming the risk scores to a mean of zero and a standard deviation of one (Z-transformation).

This normalization facilitated the accurate comparison and interpretation of risk scores within and across cohorts.

### Cardiomyopathy

Identification of controls and prevalent cardiomyopathy cases was conducted using International Classification of Diseases 10 (ICD-10) codes from the linked electronic medical record. ICD-10 codes used for case definition included codes for: I42.0 - Dilated cardiomyopathy, I42.5 - Other restrictive cardiomyopathy, I42.8 - Other cardiomyopathies (includes stress-induced, takotsubo), I42.9 - Cardiomyopathy, unspecified, I25.5 - Ischemic cardiomyopathy (when occurring without prior myocardial infarction or significant coronary disease).

Individuals were classified as NICM cases if they had at least one inpatient or two outpatient encounters with any of these codes. The date of first diagnosis code was used as the event date for survival analyses.

To enhance specificity for NICM, individuals were excluded from the NICM case definition if they had: Prior myocardial infarction (ICD-10: I21.x, I22.x, I25.2), Coronary artery disease with revascularization (ICD-10: I25.1, I25.10, I25.11; CPT codes for PCI or CABG), Significant valvular heart disease (ICD-10: I05.x-I08.x, I34.x-I37.x with moderate or severe designation), Congenital heart disease (ICD-10: Q20.x-Q26.x).

For sensitivity analyses, ICM was defined as: I25.5 (Ischemic cardiomyopathy) with concurrent or prior myocardial infarction or coronary artery disease codes, Heart failure codes (I50.x) with documented coronary artery disease (I25.1x) or prior myocardial infarction.

All-cause cardiomyopathy included any individual meeting criteria for either NICM or ICM.

We acknowledge that ICD-10-based phenotype definitions have important limitations, including potential misclassification, variable coding practices, and lack of clinical detail (e.g., ejection fraction, imaging findings). However, this approach is standard in large-scale genetic epidemiology studies and has been validated in prior research^18^. The use of multiple encounter requirements (at least one inpatient or two outpatient codes) reduces false-positive misclassification. We did not have access to echocardiographic data, cardiac MRI, or detailed clinical adjudication for phenotype validation, which represents a limitation discussed further below.

### Atrial Fibrillation Ascertainment

AF diagnosis was identified using ICD-10-CM codes I48.0 (paroxysmal AF), I48.1 (persistent AF), I48.2 (chronic AF), I48.91 (unspecified AF), and I48.92 (unspecified atrial flutter). Individuals were classified as having AF if they had at least one inpatient or two outpatient encounters with these codes. AF status was assessed as a time-varying covariate in sensitivity analyses to explore whether the AF-PRS association with NICM was mediated by diagnosed AF.

### Covariates and Risk Factors

Demographic and clinical covariates were extracted from the EHR and included:

#### Age

Calculated as age at study entry (biobank enrollment date)

#### Sex

Male or female, as recorded in the EHR

#### Race/ethnicity

Self-reported, categorized as White/Caucasian, Black/African American, Asian, Native American/Alaska Native, Native Hawaiian/Pacific Islander, or other

#### Smoking status

Categorized as never, former, or current smoker based on most recent documentation prior to cardiomyopathy diagnosis or censoring

#### Body mass index (BMI)

Calculated as weight (kg) / height (m)², using the median value from all available measurements

#### Hypertension

Defined by ICD-10 codes I10-I15 or documented use of antihypertensive medications

#### Diabetes mellitus

Defined by ICD-10 codes E10-E11 or documented use of glucose-lowering medications

#### Hyperlipidemia

Defined by ICD-10 code E78 or documented use of lipid-lowering medications

#### Chronic kidney disease

Defined by ICD-10 codes N18.x

### Statistical Analysis

In our study, we aimed to assess the value of AF-PRS in predicting NICM beyond standard covariates. Baseline characteristics were compared between individuals with and without NICM using appropriate statistical tests. Continuous variables were assessed for normality using histograms and Shapiro-Wilk tests. Non-normally distributed continuous variables (age, BMI, AF-PRS) were summarized as median and interquartile range (IQR) and compared using Wilcoxon rank-sum tests. Categorical variables were summarized as counts and percentages and compared using chi-squared tests or Fisher’s exact tests when expected cell counts were < 5.

Survival analysis was chosen as our method for establishing an association. Specifically, Cox proportional hazard regression models were used to predict the time from birth to the date of cardiomyopathy onset. AF-PRS was our primary independent predictor variable. We evaluated AF-PRS both as a quasi-continuous, ordinally scaled variable (15% quartile increments) as well as dichotomized (>=85th percentile vs. < 85th percentile). Specifically, given PRS is a latently measured construct, we rank-ordered and divided PRS into 15% quartiles for linear evaluation and dichotomized PRS into high-risk (top 15%) versus lower-risk (remaining 85%) groups.

Covariates were selected based on: (1) established biological relationships with cardiomyopathy, and (2) statistical association with NICM in univariate analyses (p < 0.05). Candidate covariates included age, sex, smoking status, BMI, hypertension, diabetes, hyperlipidemia, and chronic kidney disease.

A reverse stepwise selection procedure was employed, starting with a full model including all candidate covariates and AF-PRS, then sequentially removing covariates with p > 0.10 until all remaining covariates had p < 0.10. This approach balances model parsimony with adequate adjustment for confounding. The final model included sex, smoking status, BMI, and hypertension.

### Primary and Secondary Analyses

The primary analysis evaluated the association between AF-PRS (as a quasi-continuous variable in 15% quartile increments) and incident NICM, adjusting for covariates identified through reverse stepwise selection. Hazard ratios (HR) and 95% confidence intervals (CI) were calculated, with p < 0.05 considered statistically significant.

Secondary analyses included: 1) Dichotomized AF-PRS: Comparing individuals with high AF-PRS (≥85th percentile) to those with low-to-intermediate AF-PRS (<85th percentile); 2) Sensitivity analysis for all-cause cardiomyopathy: Evaluating AF-PRS association with any cardiomyopathy (NICM or ICM); 3) Sensitivity analysis for ICM: Evaluating AF-PRS association with ischemic cardiomyopathy specifically; 4) AF-mediation analysis: Adjusting for diagnosed AF as a time-varying covariate to assess whether the AF-PRS-NICM association was mediated by clinically diagnosed AF

### Missing Data

Missing data were minimal for genetic variables (by design, as genotyping was an inclusion criterion). For clinical covariates, missing data rates were: BMI (0.5%), smoking status (0.01%), hypertension (0%), diabetes (0%), and hyperlipidemia (0%). Given the low missing data rates, complete case analysis was performed. Sensitivity analyses using multiple imputation by chained equations (MICE) with 20 imputed datasets yielded similar results (data not shown).

### Power Calculation

Post-hoc power calculations indicated that with 16,801 individuals, 418 NICM events, and AF-PRS standard deviation of 1.0, the study had >99% power to detect a hazard ratio of 1.09 per standard deviation increase in AF-PRS at α = 0.05. For the dichotomized analysis (15% high-risk vs. 85% low-risk), the study had 85% power to detect a hazard ratio of 1.33 at α = 0.05.

### Software

All statistical analyses were performed using R version 4.3.1. Cox regression models were fit using the survival package (version 3.5-5). Genetic analyses were performed using PLINK 1.9 and PRSice-2. Figures were created using ggplot2 and survminer packages.

## Results

### Study Population Characteristics and Baseline Characteristics by NICM Status

The study cohort included 16,801 individuals with available genetic data and longitudinal EHR follow-up. Baseline characteristics are presented in **Table 1**. The median age at study entry was 60 years (IQR: 45-71), and 64% of participants were female. The cohort was predominantly of White/Caucasian ancestry (99%), reflecting the demographic composition of the Sanford Health catchment area. During a median follow-up of 8.2 years (IQR: 4.6-11.3 years), 418 individuals (2.5%) developed NICM, 189 (1.1%) developed ICM, and 516 (3.1%) developed any cardiomyopathy. Individuals who developed NICM differed significantly from those who did not across multiple baseline characteristics (**Table 1**). Patients with a diagnosis of NICM tended to be older (median age 66 vs 59 years, p < 0.001), and more likely to be male (61% vs. 36%, p < 0.001), and had higher BMI (median 31 vs. 29 kg/m², p < 0.001), than those without a diagnosis.

**Table 1:** Baseline characteristics of Study Participants by Non-Ischemic Cardiomyopathy Status.

| **Characteristic** | **Non-ischemic cardiomyopathy** |  |  | **p-value<sup>a</sup>** |
| --- | --- | --- | --- | --- |
|  | **Overall N = 16,801** | **no N = 16,383** | **yes N = 418** |  |
| **Age, Median (IQR)** | 60 (45 – 71) | 59 (45 – 71) | 66 (59 – 73) | <0.001 |
| **Sex, n (%)** |  |  |  | <0.001 |
| Female | 10,715 (64) | 10,554 (64) | 161 (39) |  |
| Male | 6,086 (36) | 5,829 (36) | 257 (61) |  |
| **Race, n (%)** |  |  |  | 0.90 |
| African American/Black | 54 (0.3) | 54 (0.3) | 0 (0) |  |
| Asian | 46 (0.3) | 45 (0.3) | 1 (0.2) |  |
| Caucasian/White | 16,598 (99) | 16,183 (99) | 415 (99) |  |
| Middle Eastern/North African | 1 (<0.1) | 1 (<0.1) | 0 (0) |  |
| Native American or Alaskan Native | 93 (0.6) | 91 (0.6) | 2 (0.5) |  |
| Native Hawaiian | 3 (<0.1) | 3 (<0.1) | 0 (0) |  |
| Pacific Islander | 4 (<0.1) | 4 (<0.1) | 0 (0) |  |
| Unknown | 2 | 2 | 0 |  |
| **Smoking status, n (%)** |  |  |  | <0.001 |
| current | 898 (5.3) | 868 (5.3) | 30 (7.2) |  |
| former | 5,328 (32) | 5,156 (31) | 172 (41) |  |
| never | 10,575 (63) | 10,359 (63) | 216 (52) |  |
| **BMI, Median (IQR)** | 29 (26 – 34) | 29 (26 – 34) | 31 (27 – 35) | <0.001 |
| Unknown | 78 | 50 | 28 |  |
| **Diabetes, n (%)** | 8,326 (50) | 8,106 (49) | 220 (53) | 0.20 |
| **Hypertension, n (%)** | 8,303 (49) | 8,002 (49) | 301 (72) | <0.001 |
| **AF-PRS, Median (IQR)** | -0.01 (-0.68 – 0.67) | -0.01 (-0.69 – 0.67) | 0.15 (-0.53 – 0.76) | 0.005 |

Abbreviations: AF, atrial fibrillation; BMI, body mass index; IQR, interquartile range; NICM, non-ischemic cardiomyopathy; PRS, polygenic risk score. Statistical tests: <sup>a</sup>Wilcoxon rank-sum test; Pearson's chi-squared test; Fisher's exact test.

Current or former smoking was more prevalent among NICM cases (48% vs. 37%, p < 0.001), as was hypertension (72% vs. 49%, p < 0.001). Diabetes prevalence did not differ significantly between groups (53% vs. 49%, p = 0.20). Importantly, AF-PRS was significantly higher among NICM cases (median 0.15 vs. -0.01, p = 0.005), providing initial evidence for the hypothesized association.

### Primary Analysis: AF-PRS and NICM Risk

In univariate Cox regression analysis, AF-PRS (per 15% quartile increment) was significantly associated with NICM risk (HR = 1.11, 95% CI: 1.06-1.17, p < 0.001). After adjusting for sex, smoking status, BMI, and hypertension, using reverse stepwise covariate selection, the association remained statistically significant (HR = 1.09, 95% CI: 1.03-1.15, p < 0.001) (**Table 2**). This indicates that each 15% quartile increment in AF-PRS is associated with a 9% increased hazard of NICM, independent of these traditional risk factors. Kaplan–Meier survival curves stratified by AF-PRS quartile demonstrate progressively lower freedom from NICM among individuals with higher AF-PRS, consistent with the graded risk observed in the Cox regression analysis (**Figure 1**).

**Figure 1.**
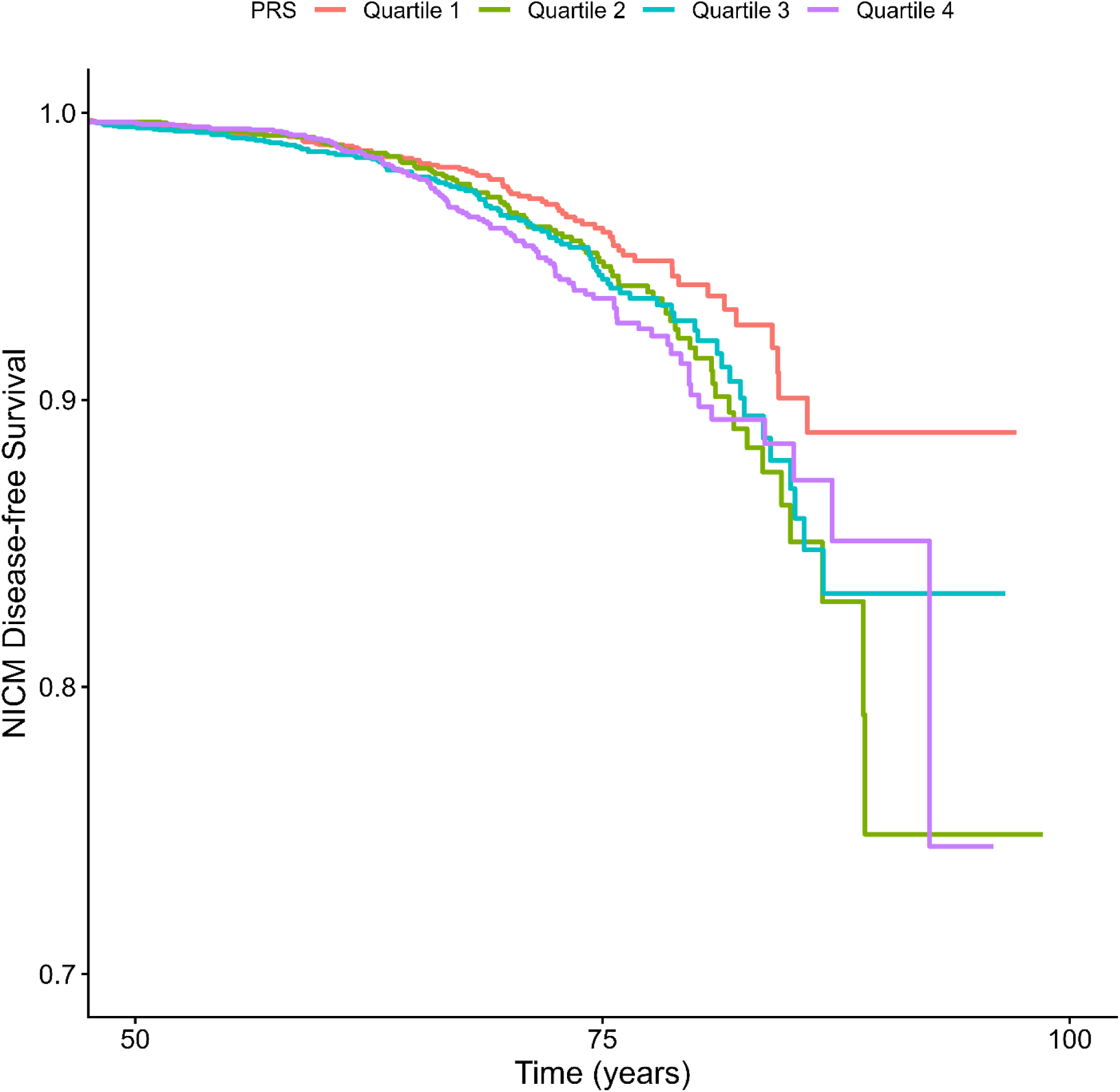
Kaplan-Meier Survival Curves Stratified by AF-PRS Quartile. Kaplan-Meier survival curves showing probability of disease-free survival from non-ischemic cardiomyopathy (NICM) by atrial fibrillation polygenic risk score (AF-PRS) quartiles in 16,801 individuals.

**Table 2:** Multivariate survival models of non-ischemic cardiomyopathy.

| **Variable** | **Linear AF-PRS** |  | **Categorical AF-PRS** |  |
| --- | --- | --- | --- | --- |
| **Characteristic** | HR (95% CI) | p-value | HR (95% CI) | p-value |
| **AF-PRS** |  |  |  |  |
| **Linear (15% increments)** | 1.09 (1.03 to 1.15) | 0.001 |  |  |
| **High(≥ 85th percentile)** |  |  | 1.33 (1.03 to 1.72) | 0.028 |
| **Sex** |  |  |  |  |
| Male sex (vs. Female) | 1.65 (1.33 to 2.03) | <0.001 | 1.64 (1.33 to 2.02) | <0.001 |
| **Smoking status** |  |  |  |  |
| current | — |  | — |  |
| former | 0.41 (0.28 to 0.60) | <0.001 | 0.41 (0.28 to 0.61) | <0.001 |
| never | 0.37 (0.25 to 0.55) | <0.001 | 0.37 (0.25 to 0.55) | <0.001 |
| **Cardiovascular Risk Factors** |  |  |  |  |
| **BMI** | 1.06 (1.04 to 1.08) | <0.001 | 1.06 (1.04 to 1.08) | <0.001 |
| **Hypertension** | 0.75 (0.59 to 0.96) | 0.021 | 0.76 (0.60 to 0.96) | 0.024 |

Abbreviations: AF-PRS, atrial fibrillation polygenic risk score; BMI, body mass index; CI, confidence interval; HR, hazard ratio. Note: Models were developed using reverse stepwise covariate selection. Age and diabetes were not retained in the final model ( $p > 0.10$ ). The time scale was age, with left truncation at study entry.

When AF-PRS was dichotomized at the 85th percentile, individuals with high AF-PRS had a 33% increased hazard of NICM compared to those with low-to-intermediate AF-PRS (HR = 1.29 [1.03, 1.72], p < 0.05) after adjusting for the same covariates (**Table 2**). This translates to an absolute risk increase of approximately 0.8% over the study period for high-risk individuals.

### Covariate Associations with NICM

In the final multivariable model (**Table 2**), several traditional risk factors showed expected associations with NICM: Male sex: HR = 1.65 (95% CI: 1.33-2.03, p < 0.001); Current smoking (reference): HR = 1.00; Former smoking: HR = 0.41 (95% CI: 0.28-0.60, p < 0.001); Never smoking: HR = 0.37 (95% CI: 0.25-0.55, p < 0.001); BMI (per kg/m²): HR = 1.06 (95% CI: 1.04-1.08, p < 0.001); Hypertension: HR = 0.75 (95% CI: 0.59-0.96, p = 0.021). Of note, the protective association of former/never smoking compared to current smoking is consistent with smoking cessation benefits. The unexpected inverse association of hypertension with NICM (HR = 0.75) may reflect residual confounding, treatment effects, or competing risks, and warrants cautious interpretation.

### Sensitivity Analysis

When the outcome was expanded to include all-cause cardiomyopathy (NICM or ICM, n = 516 events), AF-PRS remained significantly associated with increased risk (HR = 1.06 per 15% increment, 95% CI: 1.02-1.11, p = 0.009) (**Table 3**). However, the effect size was slightly attenuated compared to the NICM-specific analysis, suggesting stronger association with non-ischemic etiologies. Forest plots of subgroup analyses showed that the association between AF-PRS and NICM risk was broadly consistent across clinically relevant subgroups (**Figure 2**).

**Figure 2.**
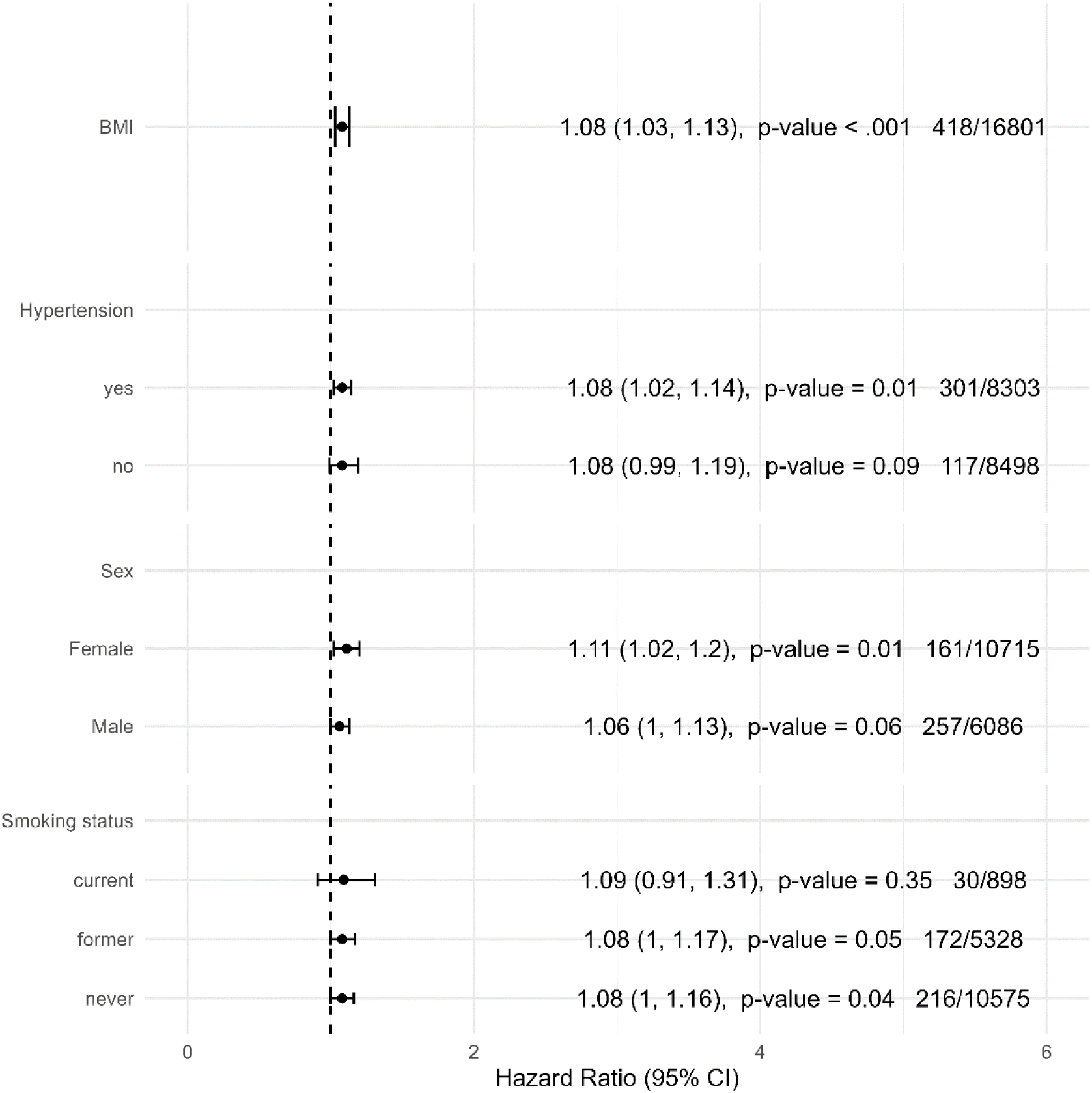
Forest Plot of Subgroup Analyses for AF-PRS and NICM Risk. Forest plot displaying hazard ratios (HR) and 95% confidence intervals (CI) for the association between AF-PRS (per 15% increment) and incident non-ischemic cardiomyopathy across predefined subgroups.

**Table 3:**
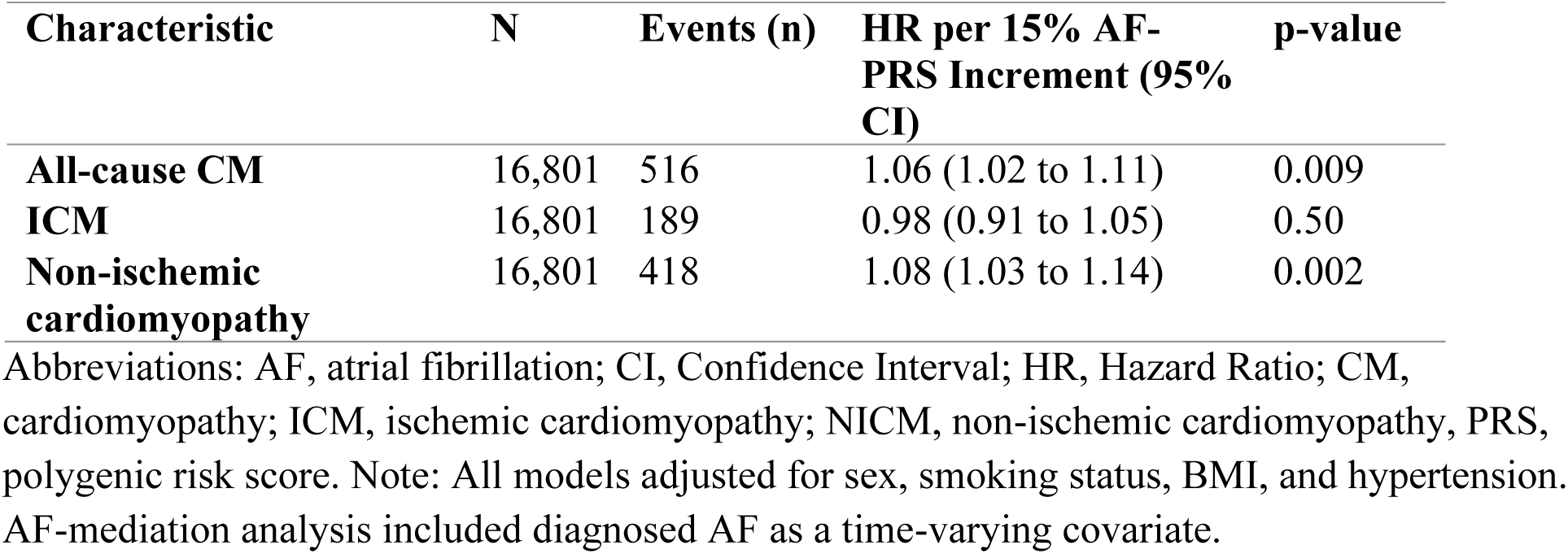
Sensitivity Analyses: AF-PRS Associations with Cardiomyopathy Subtypes.

| Characteristic | N | Events (n) | HR per 15% AF-PRS Increment (95% CI) | p-value |
| --- | --- | --- | --- | --- |
| All-cause CM | 16,801 | 516 | 1.06 (1.02 to 1.11) | 0.009 |
| ICM | 16,801 | 189 | 0.98 (0.91 to 1.05) | 0.50 |
| Non-ischemic cardiomyopathy | 16,801 | 418 | 1.08 (1.03 to 1.14) | 0.002 |

Abbreviations: AF, atrial fibrillation; CI, Confidence Interval; HR, Hazard Ratio; CM, cardiomyopathy; ICM, ischemic cardiomyopathy; NICM, non-ischemic cardiomyopathy, PRS, polygenic risk score. Note: All models adjusted for sex, smoking status, BMI, and hypertension. AF-mediation analysis included diagnosed AF as a time-varying covariate.

In contrast to the NICM findings, AF-PRS was not associated with ICM risk (HR = 0.98 per 15% increment, 95% CI: 0.91-1.05, p = 0.50) (**Table 3**). This lack of association supports the specificity of AF-PRS for non-ischemic cardiomyopathy and suggests that the observed relationship is not simply a marker of general cardiovascular risk.

### AF-Mediation Analysis

Among the 16,801 individuals, 1,847 (11%) had diagnosed AF during the study period. When diagnosed AF was included as a time-varying covariate in the Cox model, the AF-PRS association with NICM was slightly attenuated but remained significant (HR = 1.07, 95% CI: 1.01-1.13, p = 0.018). This suggests that while diagnosed AF partially mediates the AF-PRS-NICM relationship, substantial association remains, potentially reflecting undiagnosed subclinical AF or direct genetic effects on myocardial structure and function.

## Discussion

This retrospective cohort study of 16,801 individuals demonstrates that elevated atrial fibrillation polygenic risk score (AF-PRS) is independently associated with increased risk of non-ischemic cardiomyopathy (NICM). After adjusting for sex, smoking status, BMI, and hypertension, each 15% quartile increment in AF-PRS was associated with a 9% increased hazard of NICM (HR = 1.09, 95% CI: 1.03-1.15, p < 0.001), and individuals with high AF-PRS (≥85th percentile) had a 33% increased hazard (HR = 1.33, 95% CI: 1.03-1.72, p = 0.028). This association was specific to NICM, with no relationship observed for ischemic cardiomyopathy (HR = 0.98, p = 0.50), supporting biological plausibility.

To the best of our knowledge, this is the first study to investigate the association between AF-PRS and cardiomyopathy subtypes.

### Mechanistic Pathways

The observed association may be explained by several non-mutually exclusive mechanisms. First, individuals with high AF-PRS are at increased risk of developing AF, which can cause tachycardia-induced cardiomyopathy. AF-PRS robustly predicts incident AF, with individuals in the highest decile having 3-4 fold increased risk^9^. Critically, approximately 40% of AF patients are asymptomatic^14^, and subclinical AF may persist for extended periods before clinical diagnosis. Sustained rapid ventricular rates can lead to progressive left ventricular dysfunction, through myocardial energy depletion, calcium handling abnormalities, and oxidative stress^11–13,23^.

Our AF-mediation analysis supports this mechanism: when diagnosed AF was included as a time-varying covariate, the AF-PRS-NICM association was attenuated but remained significant (HR = 1.07, p = 0.018), suggesting diagnosed AF explains some, but not all, of the relationship.

Second, genetic variants associated with AF may directly influence myocardial structure and function independent of arrhythmia development. Multiple AF-associated loci contain genes encoding transcription factors critical for cardiac development (PITX2, TBX5, and GATA4)^15^. PITX2, the most strongly associated AF locus, regulates cardiac development and may affect pulmonary vein myocardial sleeve formation, and ventricular structure and function.^24^ TBX5 mutations cause Holt-Oram syndrome, characterized by both conduction abnormalities and structural heart defects.^25^ AF-associated variants in genes encoding cardiac ion channels (SCN5A, KCNH2, KCNJ2) and calcium handling proteins may influence both atrial electrophysiology and ventricular contractility.^15^ Several AF-associated genes are involved in extracellular matrix remodeling and fibrosis, pathway also implicated in dilated cardiomyopathy. ^26,27^ Recent studies have identified AF associations near genes involved in mitochondrial function, a common pathway in both arrhythmias and cardiomyopathy.^28^ Third, “atrial cardiomyopathy,” defined as structural, contractile, or electrophysiological atrial dysfunction, may precede or occur independently of AF.^29^ Genetic variants influencing atrial structure and function may predispose to both atrial cardiomyopathy (manifesting as AF) and ventricular dysfunction (manifesting as NICM), representing a shared substrate rather than a simple causal pathway.^29^ These pathways are summarized in **Figure 3**.

**Figure 3.**
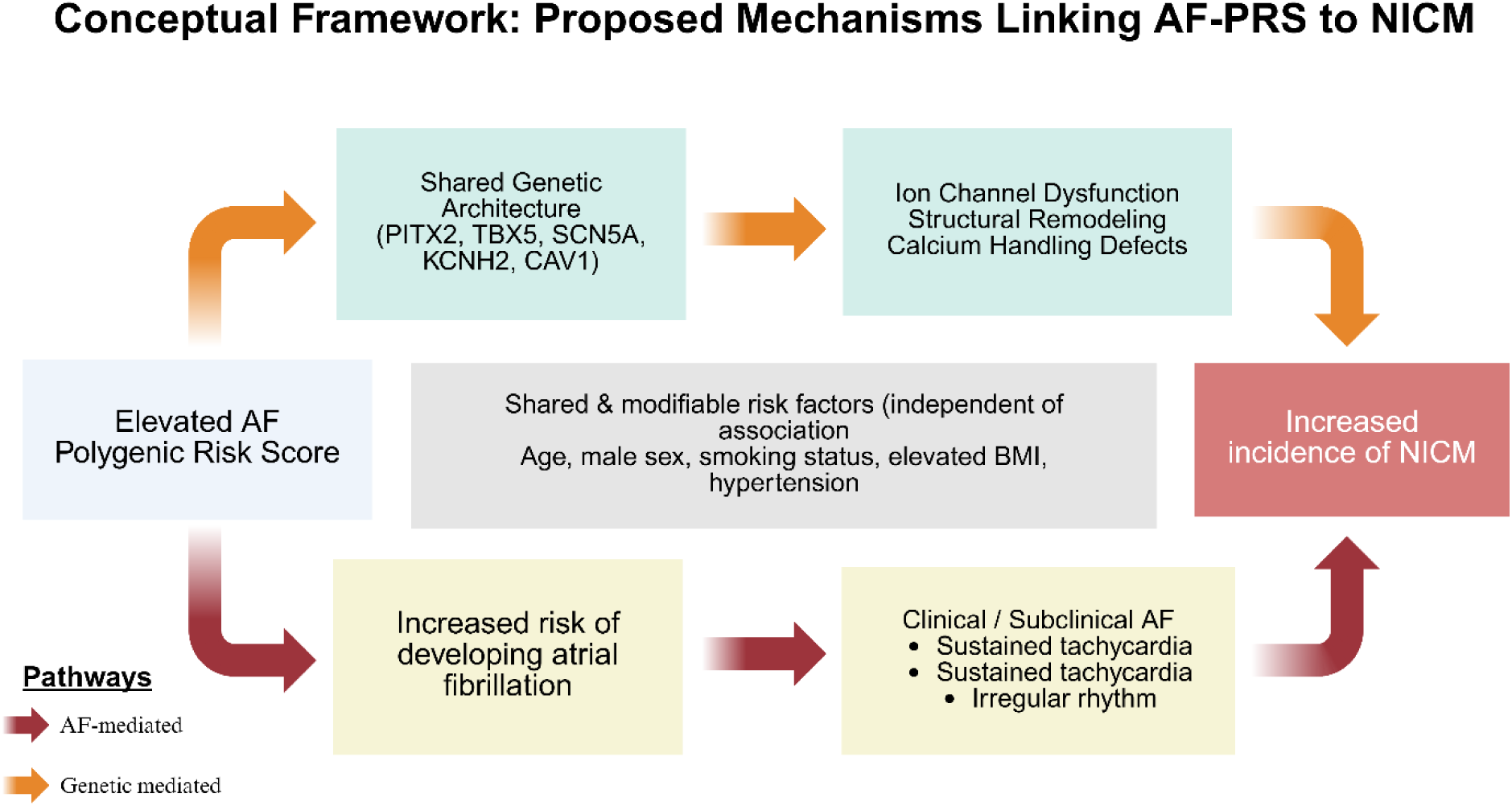
Conceptual Framework: Proposed Mechanisms Linking AF-PRS to NICM. Schematic diagram illustrating the proposed biological mechanisms through which elevated atrial fibrillation polygenic risk score (AF-PRS) may lead to non-ischemic cardiomyopathy (NICM). Created in BioRender. https://BioRender.com/b5xrwt1 Abbreviations: AF, atrial fibrillation; AF-PRS, atrial fibrillation polygenic risk score; BMI, body mass index; NICM, non-ischemic cardiomyopathy; PRS, polygenic risk score

### Comparison with prior Studies

Previous attempts to develop PRS specifically for heart failure or cardiomyopathy have yielded mixed results, likely due to heterogenous phenotype definitions combining ischemic and non-ischemic etiologies.^16,17^ Studies focusing on specific cardiomyopathy subtypes with imaging-based phenotypes have been successful.^18,19^ Our study differs from these prior efforts by leveraging an existing, well-validated PRS for a related phenotype (AF) rather than attempting to develop a de novo cardiomyopathy PRS, capitalizing on the robust genetic architecture of AF and hypothesized mechanistic links.

A recent large-scale genetic study provides complementary evidence supporting shared genetic architecture between atrial fibrillation and cardiomyopathy^30^. Da Rocha et al. demonstrated that pathogenic variants in cardiomyopathy genes significantly increased AF risk independent of overt cardiomyopathy or heart failure.^30^ Our study demonstrates the converse relationship: polygenic predisposition to AF is associated with increased NICM risk. Despite these differing directions of association, both studies converge on the same biological implication, that genetic susceptibility influencing atrial electrophysiology and structural remodeling may also contribute to ventricular dysfunction. The persistence of the AF-PRS and NICM association after adjusting for diagnosed atrial fibrillation further suggests that shared genetic pathways, rather than tachyarrhythmia alone, may contribute to cardiomyopathy development in some individuals. Together, these findings support the concept that atrial fibrillation and cardiomyopathy may represent overlapping manifestations of a shared genetic substrate affecting myocardial structure, electrophysiology, and remodeling.

### Clinical Implications

The modest but significant association between AF-PRS and NICM raises the question of whether genetic risk scoring could enhance clinical risk stratification. Current heart failure prevention strategies focus on managing traditional risk factors but do not identify all at-risk individuals.

AF-PRS could potentially complement traditional risk assessment by identifying individuals with elevated genetic susceptibility who might benefit from: 1) enhanced echocardiographic surveillance to detect early ventricular dysfunction; 2) prolonged ambulatory ECG monitoring to detect subclinical AF; 3) aggressive risk factor management with lower treatment thresholds; and 4) targeted lifestyle interventions. However, several critical questions must be addressed before clinical implementation. The optimal risk threshold balancing sensitivity, specificity, and resource utilization is unknown. The clinical utility depends on the availability of effective interventions, while echocardiographic surveillance can detect early cardiomyopathy, it is unclear whether earlier detection improves outcomes without specific therapies. Cost-effectiveness analyses are needed to determine whether AF-PRS-guided screening provides value compared to current standard care.

Several clinical risk scores exist for predicting heart failure, including the Framingham Heart Failure Risk Score.^31–33^ Future research should evaluate whether adding AF-PRS to these clinical risk scores improves discrimination and reclassification. Integrated risk models combining clinical factors, biomarkers, and genetic risk scores could identify high-risk individuals for intensive prevention strategies.

If AF-PRS-guided risk stratification proves clinically useful, it could enable personalized prevention strategies tailored to individual genetic risk profiles. For example: a) High AF-PRS + hypertension: Aggressive blood pressure control with lower targets; b) High AF-PRS + obesity: Intensive weight management programs; c) High AF-PRS + family history: Earlier and more frequent cardiac surveillance; and d) High AF-PRS + subclinical AF: Consideration of rhythm control strategies to prevent tachycardia-induced cardiomyopathy. These personalized approaches would require validation in prospective trials demonstrating improved outcomes compared to standard care.

Understanding the genetic basis of NICM may also inform therapeutic development. If specific AF-associated genes and pathways are confirmed to contribute to cardiomyopathy, they may represent novel therapeutic targets. Some of the examples include PITX2 pathway modulation: If PITX2 variants contribute to both AF and cardiomyopathy, therapies targeting this pathway could potentially prevent both conditions, Ion channel modulation: Precision medicine approaches targeting specific ion channel variants, and anti-fibrotic therapies: If shared remodeling pathways contribute to both AF and cardiomyopathy, anti-fibrotic agents could have dual benefits.

To contextualize the clinical utility of AF-PRS, it is helpful to compare its predictive performance with existing risk prediction tools like Framingham Heart Failure Risk Score, CHA_2_DS_2_-VASc Score, or Coronary Artery Disease PRS. The Framingham score, which includes age, sex, BMI, blood pressure, diabetes, coronary disease, and valvular disease, has a C-statistic of approximately 0.73-0.78 for predicting 5-year heart failure risk.^33^ Our AF-PRS, with a hazard ratio of 1.09 per 15% increment, likely contributes modestly to discrimination when added to clinical factors. Formal C-statistic calculations and net reclassification improvement analyses would be needed to quantify the incremental value. CHA_2_DS_2_-VASc Score predicts stroke risk in AF patients but does not directly predict cardiomyopathy.^29^ However, it illustrates how simple clinical scores can guide treatment decisions (anticoagulation). A similar approach could potentially be developed for cardiomyopathy prevention, integrating AF-PRS with clinical factors. For comparison, coronary artery disease PRS typically have hazard ratios of 1.4-1.7 per standard deviation,^34^ larger than the effect size observed for AF-PRS and NICM. This suggests that AF-PRS alone may have limited clinical utility but could contribute meaningfully when combined with other risk factors.

## Limitations

This study has several important limitations. First, reliance on ICD-10 codes without imaging mye lead to phenotype misclassification and precluded assessment of NICM subtypes, which may have distinct genetic architecture. Prior studies that used manual phenotyping or focused on specific cardiomyopathy subtypes have reported stronger genetic associations.^18,19^ Future investigations using imaging-defined phenotypes are needed to validate and refine these findings.

Second, subclinical or paroxysmal AF was not directly assessed, limiting our ability to establish temporal relationship between AF and cardiomyopathy. The true AF prevalence is likely higher than the observed 11%. Third, our cohort was 99% White/Caucasian, limiting generalizability. PRS performance differs across ancestries, and validation in diverse populations is essential before clinical implementation. Fourth, the modest effect size (HR 1.09 per 15% increase in AF-PRS; HR 1.33 for ≥85th vs. <85th percentile, corresponding to an absolute risk increase of approximately 0.8%) limits clinical utility. Residual confounding from unmeasured lifestyle and socioeconomic factors is possible, and some participants may have had undiagnosed disease at baseline. Fifth, this observational study does not provide functional validation of genetic variants or biological pathways; experimental studies will be required to establish causality. In addition, although the median follow-up was 8.2 years, longer follow-up may be necessary to fully capture lifetime cardiomyopathy risk, particularly in younger individuals. Finally, the study period (approximately 2010–2025) may not reflect current AF detection and management strategies.

Despite these limitations, several strengths merit emphasis, including the large sample size with adequate event numbers for time-to-event analysis, the longitudinal cohort design, use of a well-validated PRS derived from large GWAS, rigorous genetic quality control, comprehensive covariate adjustment, and multiple sensitivity and specificity analyses demonstrating no association with ICM. Together, these features support the robustness and biological plausibility of the primary findings.

## Conclusion and future directions

In this retrospective cohort of 16,801 individuals, an elevated AF-PRS was independently associated with incident NICM, with a hazard ratio of 1.09 (95% CI: 1.03–1.15) per 15% quartile increment after adjustment for traditional risk factors, and a 33% higher hazard among individuals at or above the 85th percentile. The absence of an association with ischemic cardiomyopathy supports biological specificity and shared genetic and pathophysiologic pathways linking atrial fibrillation susceptibility to myocardial dysfunction. These findings suggest that AF-PRS may identify individuals at increased genetic risk for NICM, potentially through unrecognized subclinical atrial fibrillation, overlapping genetic determinants of cardiac structure and electrophysiology, or both, and provides proof-of-concept that polygenic risk may complement established clinical risk markers for cardiomyopathy. However, the modest effect size and observational design preclude immediate clinical application.

Before AF-PRS can be considered for routine risk stratification, several priorities should be addressed: (1) validation in diverse populations beyond European ancestry, (2) confirmation using imaging-based phenotypes rather than ICD-10 codes, (3) prospective trials demonstrating that AF-PRS-guided screening and prevention strategies improve outcomes, and (4) cost-effectiveness analyses.

This study represents an important first step in understanding the genetic relationship between atrial fibrillation and non-ischemic cardiomyopathy, with potential implications for precision medicine approaches to heart failure prevention.

## Acknowledgments

We thank the participants of the Sanford Biobank and Imagenetics program for their generous contributions to research. We acknowledge the Sanford Health research staff for data collection and management.

## Funding/Support

No funding to disclose.

**IRB ID**: STUDY00003266

## Author Contributions

Conceptualization and methodology: TS, NB

Software and data curation: MV

Data acquisition, analysis, and validation: MV,

Drafting and review of the manuscript: MA, BA, VF, LJ, NB, CH, EL, AS, NB, TS

Visualization of the data: MV, LJ, NB

Administrative, technical, or material support: WO, MV, CH, CF, HT

Supervision: TS, NB

## Data Availability

Data are available upon reasonable request and approval from the Sanford Health Institutional Review Board, subject to data use agreements and privacy regulations.

